# Readiness Among Female Community Health Volunteers to Deliver Community Based Tobacco Cessation Counseling in Rural Nepal: A Two-phase Mixed-methods Study

**DOI:** 10.64898/2026.11.10.26367774

**Authors:** Omkar Jha, Basanta Chaulagain, Susagya Bhusal, Ada Thapa, Sudim Sharma, Prayas Gautam, Austin Seals, Adipti Pantha, Manoj Bhattarai, Bibhav Acharya, Yashashwi Pokharel

## Abstract

**Background:** It is unknown whether community health workers called Female Community Health Volunteers (FCHVs) can deliver tobacco cessation counseling in Nepal.

**Methods:** Using the Health Belief Model, we conducted a two-phase mixed-methods evaluation among 28 FCHVs in rural mid-Western Nepal in eight Focused Group Discussions (FGDs) exploring FCHVs’ knowledge, self-efficacy, and barriers to provide tobacco cessation counseling in phases 1 and 2 eleven months apart. After phase 1 FGDs, we conducted a training about tobacco cessation using didactics and role play and a refresher training after phase 2 FGDs. In each phase, we concurrently administered two surveys before and after each training. We utilized deductive and inductive codes in qualitative analysis and identified themes from the codes. We used descriptive statistics and Fisher’s exact test for quantitative data, and concurrent mixed methods (Qual + quan) during data interpretation for convergence and complementarity.

**Results:** FCHV’s mean age was 48 ± 6.8 years, 1/3^rd^ had secondary education, and 3/4^th^ was working as FCHVs for >10 years. Qualitative analysis revealed five themes about FCHVs’ knowledge, self-efficacy and barriers. During phase 1, FCHVs demonstrated a good knowledge of tobacco, its effects on health, and tobacco cessation measures. They recognized having a role in cessation counseling but stressed needing regular trainings. In phase 2, while some misconceptions remained, FCHVs’ knowledge and confidence improved, including their trust in the achievability of tobacco cessation. However, hesitation persisted. They identified several personal, social and policy-level barriers. Communities’ receptivity to counseling improved in phase 2, however the COVID pandemic disrupted their efforts. Quantitative analysis revealed that FCHVs not only identified having a role, but also routinely provided counseling. There was relative stability of the baseline knowledge, confidence and perceived barriers across surveys. In mixed methods evaluation, information converged but also complemented. Qualitative inquiries provided depth and quantitative evaluation showed numerical trend.

**Conclusion:** FCHVs want to provide tobacco cessation counseling, had good knowledge and counseling skills, but faced several barriers. These improved after training. Policy makers should consider addressing other system-level barriers. Future studies should evaluate the clinical effectiveness of this approach before adoption in routine practice.

## Background

Tobacco use is responsible for over 8 million deaths annually, including 1.3 million from second hand smoke exposure [1]. The burden of tobacco-related illness is especially pronounced in low and middle-income countries (LMICs), where approximately 80% of tobacco users reside [1]. In Nepal, the prevalence of tobacco consumption persists at alarming rates of 28.9% (48.3% in males and 11.6% in females) [2], and the tobacco-related deaths is 14.9% of yearly fatalities [3]. Broadly accessible tobacco cessation strategies are urgently needed in Nepal.

Tobacco cessation counseling by health care providers is an evidence-based approach [4]. However, access to effective cessation services is very limited with only one-third of the world’s population having access to programs meeting best practice standards [1]. A community-based scalable tobacco cessation counseling approach can meet this unmet need.

Tobacco cessation support is notably lacking in healthcare establishments in Nepal [2]. Additionally, medications, including Nicotine Replacement Therapies (NRTs), are not readily available in rural areas, even though the World Health Organization lists NRT as an essential medication [5]. Community health workers (CHW) commonly called Female Community Health Volunteers (FCHV) in Nepal, represent their community, are well respected, and have demonstrated their ability to manage other non-communicable diseases [6, 7]. Thus, FCHV can have an important role in community-based tobacco cessation counseling to combat tobacco related illness in rural Nepal and can be a model for other countries. We explored the potential for FCHVs to provide tobacco cessation counselling in rural Nepal.

## Methods

### Study setting and population

This study was part of the Non-Communicable Disease (NCD) in Nepal Study, which uses community-based interventions to address NCD in rural Nepal in four administrative divisions known as wards within the Ghorahi sub-metropolitan city [8]. Each ward is a local administrative unit responsible for various community-level services. The four wards have a total population of 31,840 residents and 45 FCHVs. We contacted all 45 FCHVs, and 28 agreed to participate.

### Study design

We used a two-phase concurrent embedded mixed-method design for data collection and evaluation (Qual + quan, Figure 1) [9]. The Health Belief Model (HBM) informed our semi-structured interview guides and quantitative survey questions [10, 11]. The model comprises six socio-behavioral constructs: perceived benefits, barriers, susceptibility, severity, the presence of cues to action, and self-efficacy that could shape health behaviors, often interacting with other intrapersonal factors [10, 11]. For our study, we focused on FCHV’s knowledge, self-efficacy, and barriers to provide tobacco cessation services. These constructs are relevant in assessing how FCHVs perceive and manage tobacco cessation counseling, as they reflect the internal and external factors that influence their counseling practices. By applying the HBM, we aimed to capture how FCHVs’ beliefs about the effectiveness and challenges of tobacco cessation counseling shape their attitudes and approaches to delivering these services [10, 11]. Each phase included four Focus Group Discussions (FGDs), a survey before a training on tobacco cessation measures, and a survey after the training (Figure 1). In total, there were four surveys and two rounds of FGDs. We spent ∼7-8 hours per ward delivering the FGDs, surveys, and training during each phase. The program’s broader goal was to assess FCHVs’ needs and readiness to address hypertension, diabetes, hyperlipidemia, and tobacco cessation in their communities. The current study is focused on tobacco cessation. We offered light refreshments and provided a tote bag and a modest reimbursement (US ∼$4 per phase) to each FCHV following local norms.

**Figure 1:**
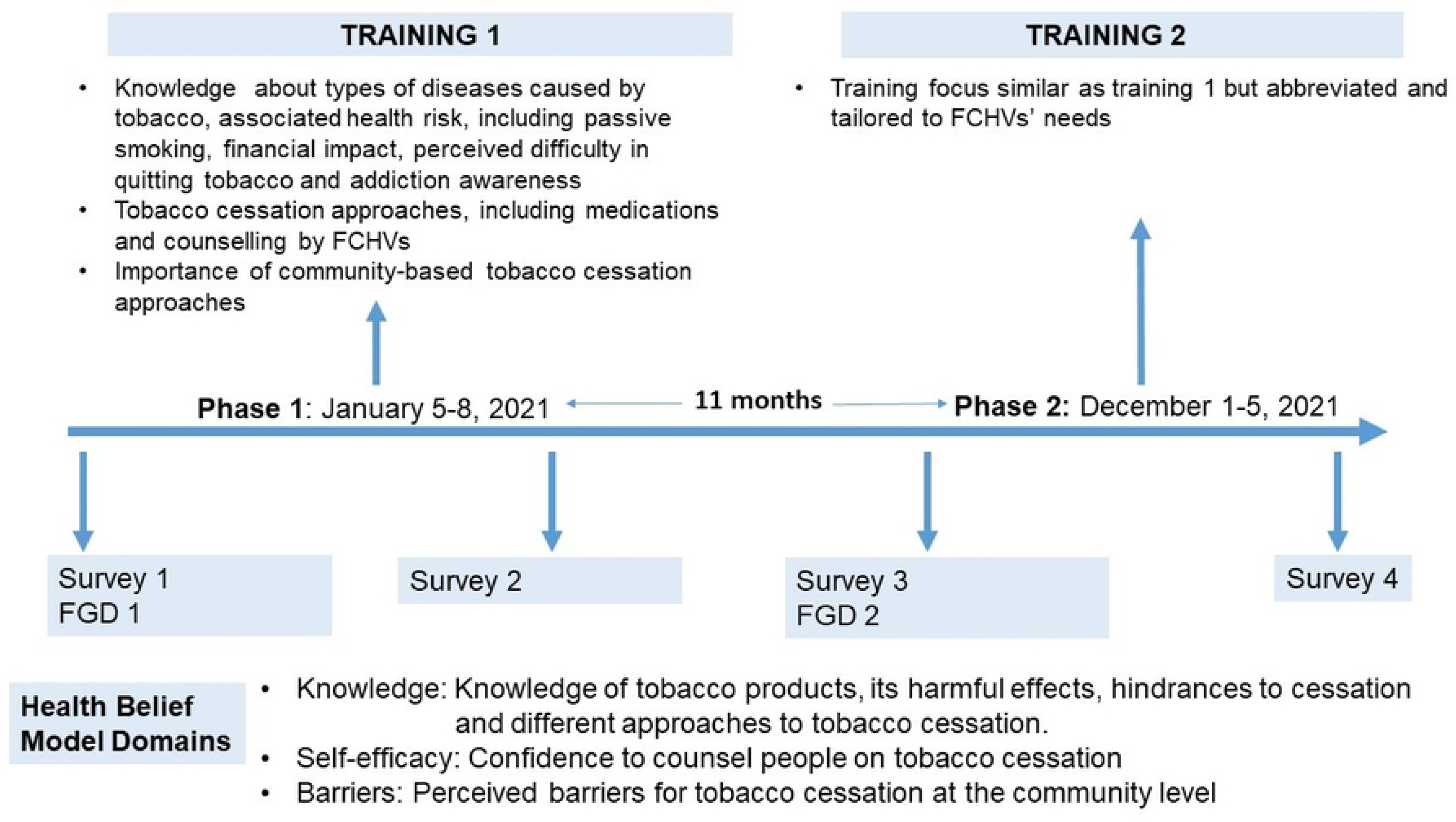
Study timeline and relevant Health Belief Model domains Legend: FCHV: Female Community Health Volunteer, FGD: Focused Group Discussion

### Data collection instruments

The semi-structured interviews conducted during FGDs provided rich insights into FCHVs’ knowledge of the health risks associated with tobacco use, their self-efficacy in delivering cessation counseling, and the barriers they face in implementing such interventions. These interviews allowed us to explore the contextual and experiential factors influencing FCHVs’ engagement with tobacco cessation efforts, offering a nuanced understanding that complemented numerical data. To further examine these domains, we also evaluated FCHVs’ understanding, confidence, and challenges using quantitative surveys, enabling a more comprehensive assessment of their perspectives and practices (Supplemental file).

Two people independently translated the interview guides and the survey questions developed initially in English into Nepali language. We reconciled the translation to ensure linguistic accuracy and cultural relevance through team discussions. We pretested the study instruments in a small sample of people similar to the target population. Feedback from the pretesting improved clarity, relevance, and appropriateness of the study instruments.

We collected both qualitative and quantitative data simultaneously but independently across two phases (Figure 1). Phase 1 took place from January 5, 2021, to January 8, 2021, and phase 2 took place from October 9, 2021, to October 12, 2021.

### Phase 1

#### Survey 1

We delivered a survey comprising nine questions exploring FCHVs’ knowledge, barriers and skills in providing tobacco cessation counseling (Supplemental file, Figure 1).

#### FGD 1

Prior to the initial training, we conducted one FGD per ward with seven FCHVs participating per FGD. Each FGD lasted approximately 45 minutes. The NCD Nepal Study staff who have previously interacted with some FCHVs conducted the semi-structured interviews in local public schools and health facilities.

#### Training 1

We conducted a structured training program aimed to enhance FCHVs’ knowledge and skills to deliver impactful smoking cessation counseling covering topics ranging from the health risks of smoking to counseling techniques and community-based approaches.

Each training session lasted for ∼4 hours and included:

- Didactic sessions delivered by physicians on topics like health risks of smoking and counseling techniques.
- A variety of learning methods, including presentations, posters, pamphlets and interactive activities.
- Role-play demonstrations to practice counseling skills using the 5A’s brief intervention, which includes asking about smoking status, assessing readiness to quit, advising to quit, assisting with the quitting process, and arranging follow-up counseling. Evidence shows that the 5A’s brief intervention can increase quit rates among smokers [12]. We also discussed about the principles of motivational interviewing and shared decision making.

#### Survey 2

Following the initial training, we administered a survey to assess any immediate changes in FCHVs’ knowledge and skills. Survey 2 had seven tobacco cessation related questions like survey 1 (survey 2 aimed to gauge the effectiveness of the training; two questions from survey 1 were removed, see Supplemental file).

### Phase 2

After 11-months from phase 1, we evaluated retention of knowledge and skills, FCHVs’ experience and challenges in delivering tobacco cessation counseling in the community using interview guides and survey tools like phase 1 (Supplemental file). We started with FGD2, survey 3, a refresher training (training 2) and survey 4 following the training (Figure 1). The refresher training was similar to phase 1, but was also tailored to overcome challenges encountered in delivering effective cessation counseling in the last 11 months.

### Data analysis

#### Qualitative data analysis

We audio recorded FGDs. Research staff transcribed the interviews and cross-checked for accuracy. We prioritized information power during data collection over thematic saturation and used both deductive and inductive codes to identify themes [13, 14]. Pre-established codes from the Health Belief Model informed deductive coding, while inductive coding allowed emergence of new information from the data [15]. A coding team comprised of 3 trained staff developed and iteratively revised the codes from eight transcripts (one from each FGD) reaching a consensus by discussion among the coding team and with the study principal investigator and generated the final codebook. The coding team subsequently applied the codebook to all the transcripts using Atlas.ti software. We subsequently generated code report for each code. We then developed a code summary which informed development of themes based on the patterns and relationships among the codes after discussing with the investigative team.

#### Quantitative data analysis

We analyzed quantitative data in R (4.0.2) using descriptive statistics for socio-demographic variables and Fisher’s exact test for smaller sample group. We presented descriptive results as mean, standard deviations (SD), and frequencies.

To evaluate FCHVs’ training needs, we pre-specified comparisons of results between surveys 1 vs. 2 to evaluate immediate change in knowledge after the training, surveys 2 vs. 3 to evaluate short term retention of knowledge, and surveys 1 vs. 4 to evaluate the overall change.

### Integration of data

Our study involved independently but concurrently collected and analyzed quantitative data within the qualitative dominant evaluation (Qual + quan, Figure 2) [9]. We integrated the results during interpretation for convergence and complementarity [9]. Quantitative analysis provided numerical observed trends corroborating key themes and patterns from qualitative data. Qualitative insights highlighted the nuances and context specific information influencing FCHVs’ engagement with smoking cessation counseling, adding depth to the quantitative results.

**Figure 2:**
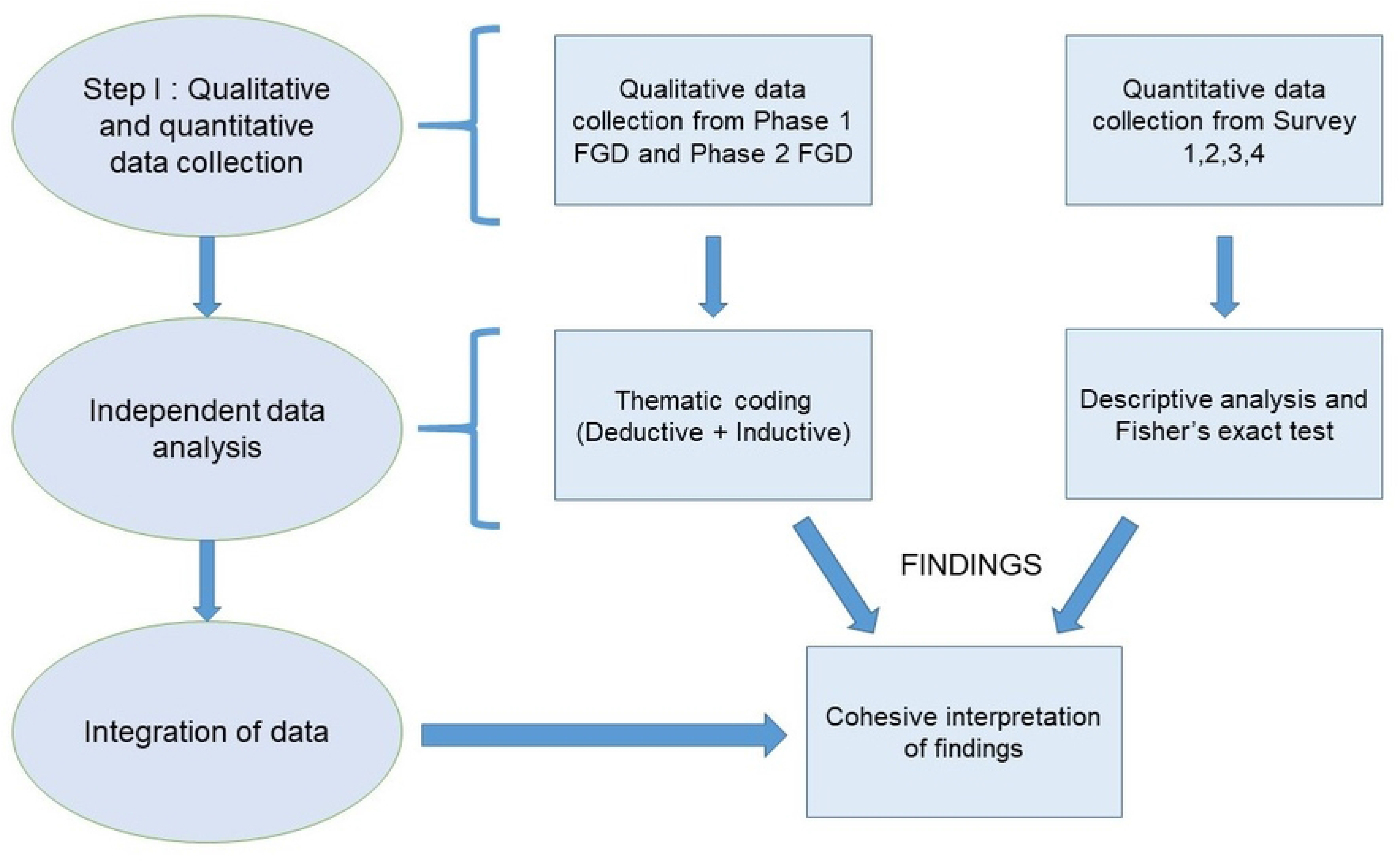
Integration of qualitative and quantitative data Legend: FGD: Focused Group Discussion

### Ethical consideration

The research activities for this study were carried out in accordance with the Declaration of Helsinki. This study is part of an ongoing NCD Nepal Study [8]. We obtained ethical approval from Nepal Health Research Council and a written consent from each FCHV. Before obtaining consent, data collectors informed participants that their participation was voluntary, explained the purpose of the study and the interview, and discussed logistics, the use of audio recording, and de-identification procedures.

## RESULTS

### Sociodemographic data

FCHVs’ mean age was 48 ± 6.8 years, 9 had secondary education and 21 had more than >10 years of experience working as an FCHV (Table 1).

### Main findings

We found five themes in qualitative analysis.

**1) There was a strong baseline understanding among FCHVs regarding the health and social effects of tobacco, which improved after training, though some misconceptions persisted.**

In Phase 1, FCHVs were able to list a range of tobacco products, including cigarettes, *bidis*, and chewing tobacco, with misconception like alcohol as a type of smoking product. FCHVs agreed that smoking has *harmful health effects, particularly on the lungs, heart, liver, kidneys* and causes various types of cancer. We noted misunderstanding like smoking causing tuberculosis. *The harmful effects extended beyond the smoker*, with some FCHVs recognizing the dangers of secondhand smoke. One participant noted the far-reaching impact, saying,

> Quote 1: *"Tobacco use includes smoking cigarettes and drinking home-brewed alcohol. Tobacco affects the lungs, causes heart disease, and leads to high blood pressure. It can also cause tuberculosis. Even if you one person smoke, it will affect all your family members."* –FGD 4, Ward No. 4, Phase 1

By Phase 2, FCHVs demonstrated a more detailed understanding of tobacco products, and were able to identify various types, such as cigarettes, *gutka* (betel quid), *bidis*, and *bhaang (*cannabis leaves). FCHVs continued to emphasize the severe consequences of smoking, and noting that *tobacco can cause different types of cancer, particularly lung and throat cancer*. One participant elaborated by saying,

> Quote 2: *"Smoking refers to tobacco-related products, such as cigarettes, gutka (betel quid), bidis, bhaang (cannabis leaves), and many others. If you use them, they will affect your lungs, turning them black, damage your heart and kidneys, and increases blood pressure. They can cause lung and throat cancer. They not only harm the smokers but also affect those who are around them."* - FGD 2, Ward No. 6, Phase 2

The survey data showed a good understanding about the harmful effects of smoking at baseline that remained consistent over time (Table 2).

Another aspect highlighted by FCHVs was the social and familial effects of smoking. In Phase 1, they acknowledged that smoking could negatively impact the family and society as a whole, though this sentiment was not as widely expressed. By Phase 2, the recognition of smoking’s broader social consequences was more pronounced, with FCHVs even pointing out that *smoking could be a reason for divorce*. Additionally, they noted the positive impact of quitting tobacco on family dynamics, emphasizing improvements in relationships.

In Phase 1, FCHVs noted the harmful effects of smoking on the unborn child, expressing *it was particularly important to counsel pregnant women to avoid smoking*. In Phase 2, this recognition was more evident. One participant shared,

> *Quote 3: "We do counsel. We counsel pregnant women not to smoke. If a pregnant women smokes, it not only affects the pregnant woman, but also the unborn child in the stomach."* – FGD 3, Ward No, 3, Phase 2

When integrating qualitative and quantitative data together, we noted that although FCHVs had a good baseline knowledge of tobacco products and their harmful effects, there were nuanced differences and the depth of understanding. Serial surveys did not show significant changes, but when considered together with qualitative results, we noted improved knowledge in second phase, underscoring the importance of education (Table 3).

**2) Easy access to tobacco, observational learning, peer influence, perceived coping mechanism for stress were considered relevant for tobacco initiation and as hindrances to overcome the addictive nature of tobacco.**

In Phase 1, FCHVs identified the easy availability of tobacco products as a significant factor hindering smoking cessation. They emphasized that *tobacco products are widely available*, and noted that *tobacco is very cheap*. They suggested that *the sales of tobacco should be restricted to specific areas*, and recommended implementing age restrictions on tobacco sales to prevent easy access to children. FCHVs also proposed making tobacco products more expensive to reduce their accessibility. A participant in Phase 1 explained,

> Quote 4: "*Whichever store you go to, the first thing you see is tobacco and cigarettes. There should be a specific place to purchase products like tobacco. Tobacco products shouldn’t be available to be purchased everywhere and there should be a specific time to buy them.*" – FGD 3, Ward No. 3, Phase 1

By Phase 2, FCHVs continued to highlight the availability of tobacco as a major barrier to cessation and recommending specific restrictions on where tobacco products should be sold. A participant stressed the importance of regulations to prevent tobacco sales near schools and other public areas,

> Quote 5: "*Importantly, to break the addiction, there should be restriction to sell tobacco products near ward offices, schools and health centers. First and foremost, the government themselves should create a system to prohibit the sale of tobacco products within certain kilometers from schools.*" – FGD 3, Ward No. 3, Phase 2

FCHVs in both Phase 1 and Phase 2 identified several factors influencing the initiation of smoking, with a strong emphasis on early exposure and peer pressure. FCHVs noted that *children often learn smoking by observing elderly family members* or when children are sent to buy cigarettes. Peer pressure was also a key factor, with FCHVs highlighting how *friends encourage each other to smoke*, which is difficult to resist. Additionally, smoking was recognized as a coping mechanism, for *dealing with stress, whether from family issues or work-related pressures* in older adults. Others associated tobacco as a helping mechanism to fall asleep. In both phases, FCHVs acknowledged that multiple factors play an important role in the *addictive nature to tobacco, making it very difficult for people quit*. They further added that withdrawal symptoms, along with long-term tobacco use, hamper cessation efforts. As one participant in Phase 1 stated,

> Quote 6: "*Small children learn smoking by watching you smoke, thinking if my father smokes cigarettes, then why shouldn’t we smoke. Others fall into the habit of smoking due to the influence of their friends and then become addicted to it. People nowadays have work pressure and family stress, which leads to mental stress and anxiety, and they smoke cigarettes as a stress reliever*." – FGD 1, Ward No. 7, Phase 1

Similarly, another FCHV in Phase 2 emphasized how peer pressure and stress continue to drive smoking behavior, saying,

> Quote 7: "*Boys often smoke with their friends and say that their friends pressure them to smoke even when they try to resist, making it hard for them to quit. Older adults claim that smoking during periods of stress helps them feel better*." – FGD 4, Ward No. 4, Phase 2

Surveys complemented the qualitative findings about the addiction nature of tobacco with high levels of agreement on needing multiple attempts to quit smoking (Table 2). The integration of qualitative and quantitative data portrayed the multifaceted factors relevant in tobacco cessation, influenced by accessibility, early habits, peer influence, addiction, and coping mechanisms (Table 3).

**3) After the training, there was a noticeable shift in FCHVs’ belief in the achievability of quitting tobacco, with multifaceted strategies such as substitutes, family support, counseling, and medical assistance.**

FCHVs in both phases expressed mixed views on the achievability of smoking cessation. In Phase 1, while there were some doubts, FCHVs also believed in the achievability of smoking cessation through personal effort or the use of medication. FCHVs also acknowledged that while smoking cessation might be achievable, *many smokers return to their old habits and start smoking again after a period of time*. In Phase 2, there was a notable shift in the belief of achieving smoking cessation, while doubts remained. FCHVs felt that smoking cessation could be achieved with the right effort, determination, support and medication. They expressed that *younger people might have a better chance of quitting, while older individuals might face more challenges*. One FCHV shared,

> Quote 8: “*It isn’t possible to quit smoking immediately. For a person to quit, it may take almost 6 months to a year.*” – FGD 3, Ward No. 3, Phase 2

FCHVs in both phases identified various smoking cessation aids. Across both phases, they suggested that keeping locally available aids, such as *Sichuan pepper, cloves, or cardamom, in the mouth could help smokers gradually reduce their dependence on tobacco*. Policy changes, such as stricter regulations on tobacco sales, were considered highly important. Counseling and support from family members were also highlighted as significant contributors to quitting smoking. Rehabilitation centers and internal motivation to quit were identified as important in both phases, while FCHVs in Phase 2 additionally recognized the roles of teachers, advice from doctors, and religious practice such as *resolution for tobacco cessation during religious pilgrimages*. One participant from Phase 1 shared,

> Quote 9: "*I have not heard of any medicine that helps to quit smoking. However, you can quit by keeping Sichuan pepper, cloves, and cardamom in your mouth and gradually reduce use of cigarettes*." – FGD 1, Ward No. 7, Phase 1

There was a notable increase in the awareness of both Nicotine Replacement Therapy (NRT) and other medications for tobacco cessation from Phase 1 to Phase 2. Additionally in Phase 1, FCHVs noted that smoking cessation achieved through medication might not last, as *individuals usually relapse once they stopped taking the medicine*. Others expressed that *people may stop taking medications due to side effects*. By Phase 2, not only FCHVs recognized the utility of medications for smoking cessation, they also cited the role of *physicians as very important for providing counseling and support in the cessation process*. However, lack of awareness or doubts about the benefits of medications were still noted. One FCHV in Phase 2 reflected,

> Quote 10: “*Smoking addiction can be addressed with the use of medicine and with rehabilitation. If all family members support and advise without scolding, then they might quit smoking by gradually decreasing tobacco use*.” – FGD 2, Ward No. 6, Phase 2

**4) FCHVs recognized their role in providing counseling for tobacco cessation, and their confidence could be further strengthened through regular training and observation of expert-led counseling sessions.**

FCHVs in both phases demonstrated confidence in their ability to counsel individuals on tobacco cessation, considering a wide range of counseling topics. In both phases, they emphasized the harmful health effects of smoking, particularly its impact on the smoker’s lungs, heart, and potential risks during pregnancy. They also highlighted the negative effects of smoking on family members, as well as its financial implications. By phase 2, they had provided counseling to community members on various strategies to reduce tobacco consumption. They remarked that *it is difficult to quit tobacco all at once, but with determination it can be achieved by gradually reducing the number of cigarettes day by day*. Additionally, FCHVs also counseled on how medications could help quit smoking. However, they felt they *lacked the necessary skills and confidence to counsel individuals who were highly regarded in the community*, such as elderly person, who in Nepali culture are respected by their youngers. One FCHV reflected,

> Quote 11: “*We have hope now to counsel more effectively than before. We advised them that smoking can lead to diseases, including cancer. Some listen to our advice to not smoke, but others mention that quitting has become difficult due to habit. We advise them, but we also need counselors* (in the community for further support).” – FGD 1, Ward No. 7, Phase 2

When evaluated across surveys, all FCHVs not only identified having a role in smoking cessation counseling but also routinely counseled. Furthermore, they expressed high confidence in counseling that remained consistent across the surveys (Table 2). In Phase 1, FCHVs expressed a desire for regular training with repeated sessions to improve their counseling effectiveness. They also suggested *including community members in training sessions* so that the community could recognize that FCHVs had received proper training to provide effective counseling. A participant articulated,

> Quote 12: *“We advise people not to smoke because it is harmful to their health. Don’t smoke as smoking affects lungs, heart, and kidneys, and can cause cancer. It would be easier for us to counsel if there is related training. A short, one-day training would not be sufficient, and we could forget; we need trainings at regular intervals.”* – FGD 3, Ward No. 3, Phase 1

By Phase 2, in addition to the need for regular training, FCHVs also highlighted the importance of *involving expert counselors* to handle cases of severe tobacco addiction. They remarked that it would be easier for them to learn *if they could observe counseling provided by experts*. They suggested that additional resources, *such as pamphlets*, could support their counseling efforts and help them *deliver more effective messages to individuals and families*.

Mixed-methods interpretation showed convergence in that FCHVs were confident in their counseling skills but complementarity by showing the nuances in qualitative inquiry (e.g., observation of counseling by a more skilled person) and in quantitative approach in that all FCHVs were already routinely counseling (Tables 2-3).

**5) Several personal, family, cultural and community level barriers in counseling were identified that improved somewhat in phase 2, while COVID pandemic related barrier emerged.**

FCHVs encountered several barriers to smoking cessation counseling, both before and after the training sessions, which affected their ability to effectively engage with the community. In Phase 1, FCHVs identified community’s receptivity to counseling as a barrier, upon which FCHVs reflected saying tobacco addiction is *a very difficult habit,* portraying tobacco use as a deeply ingrained addiction. FCHVs expressed that some community members reacted dismissively or even angrily during counseling efforts. One FCHV recounted,

> Quote 13: “*When we try to counsel, some people get angry and say, ‘Did you give me the money? I have smoked with my own money, not your father’s money.’ We don’t know much about this and lack appropriate skills, which makes it challenging for us to work in the community. Training in this area would make it easier for us to counsel*.” – FGD 4, Ward No. 4, Phase 1

FCHVs also noted that insufficient training, along with limited knowledge and skills, were significant barriers to providing effective counseling. Additionally, they identified a lack of time as a challenge, as they are *responsible for household chores and involved in various other community health projects*, which limited their availability for smoking cessation counseling. In addition, they faced challenges in reaching individuals living far away and in counseling older community members due to concern for not being respectful. FCHVs felt they *lacked the confidence to counsel elder individuals who are highly respected in the community* (in Nepali culture).

In Phase 2 after the training, some barriers persisted, but FCHVs noted improvements in community’s receptiveness to counseling, with *more community members understanding the harmful effects of smoking*. However, barriers persisted, particularly among individuals who dismissed their advice, as illustrated by one participant,

> Quote 14: “*When we counsel them to quit smoking, some respond with, ‘Did you buy this for me? Did I smoke using your money? I enjoy smoking, so I do it.’ These are some of the barriers we face. One or two people appreciate our counseling. We couldn’t provide as much counseling this time from COVID*.” – FGD 3, Ward No. 3, Phase 2

Citing the same challenges of respect and confidence when counseling older community members in Phase 2, the barrier was less prominent, as one participant reflected,

> Quote 15: “*We did not face many barriers. Some individuals now understand that smoking is harmful. There are few people nowadays (who don’t listen to us).”*– FGD 4, Ward No. 4, Phase 2

Several community members interpreted FCHVs’ counseling as a genuine concern for the community’s well-being, with a growing acceptance and appreciation of the FCHVs’ role. One FCHV shared,

> Quote 16: “*There are many people who have said that you have given good advice, which has benefited us and improved our health*.” – FGD 2, Ward No. 6.

New barriers emerged during Phase 2, including the impact of the COVID-19 pandemic, which limited counseling activities. FCHVs also cited geographical barriers to reach certain communities. To overcome the barriers, FCHVs expressed a need for ongoing training and tailored strategies to address the cultural dynamics and to improve their engagement with the community.

Results were similar across surveys (Table 2). Concerns about inadequate incentives and motivation increased from 25% to 57.1% in phases 1 to 2.

In mixed-method interpretation, we found convergence (Table 3). However, qualitative data showed emergence of new barrier (e.g., disturbance from the pandemic).

## Discussion

We evaluated whether FCHVs can deliver tobacco cessation counseling in rural Nepal. Through quantitative surveys, qualitative inquiries and their integration, we identified five themes. FCHVs demonstrated a good understanding of tobacco products and recognized both health and social ramifications of tobacco use. They identified factors relevant in tobacco initiation and cessation like easy availability of tobacco products, early experiences, peer influence, addiction, coping mechanisms and perceived stress relief from tobacco use. After the initial training, FCHVs increasingly recognized the achievability of tobacco cessation and the role of multifaceted cessation strategies including family support, counseling, policy changes, substitutes and medications. FCHVs confidence in counseling improved with the training. FCHVs expressed multi-dimensional barriers like community member’s hesitation to participate in counseling, insufficient training, inadequate time, and other challenges, but post-training showed improvements. The shift in communities’ perceptions over time indicate growing acceptance of FCHVs’ role in tobacco cessation. Even a brief and an infrequent training like ours can enhance FCHVs’ knowledge, skills, and confidence in tobacco cessation. Empowering FCHVs with comprehensive, tailored training and sustained support can enhance their effectiveness in tobacco cessation counseling.

In Nepal, access to healthcare services, especially in rural areas, remains a challenge. There is a notable absence of tobacco cessation support in healthcare facilities even in the tertiary care centers [2]. Therefore, there is a critical need to develop a scalable capacity to support tobacco cessation efforts in Nepal. FCHVs have demonstrated effectiveness in community-based care for chronic conditions like hypertension and diabetes.[7, 16] Our study shows that FCHVs are able to provide tobacco cessation counseling and have a massive scalable potential due to their presence throughout Nepal.

Our findings highlight the need for continuous education and support for FCHVs to sustain skills in smoking cessation counseling. Strengthening their skills through ongoing training and supervision enhances their ability to provide counseling services, extending the reach and impact of these initiatives. Cultivating supportive community environments that facilitate community members’ attitudes and receptivity towards cessation counseling will be helpful, such as leveraging civic groups like Mother’s Groups for advocacy with whom FCHVs closely work [17]. Integrating smoking cessation services by FCHVs into Nepal’s health policy is important. Policymakers can allocate resources for comprehensive training of FCHVs and establish referral systems to connect individuals requiring extra support. Moreover, addressing systemic barriers like tobacco marketing and accessibility through robust tobacco control measures is imperative to reduce tobacco-related health burdens in Nepal. FCHVs noted, "*If the tobacco products are made expensive, then maybe they wouldn’t buy it,"* highlighting the potential impact of taxation and pricing policies. They also pointed out the pervasive visibility of tobacco products in shops, indicating a need for stricter implementation of restriction related to age, time, and places for selling and using tobacco products.

In Nepal, access to pharmacological aids for smoking cessation, including NRTs and non-NRTs like bupropion, is limited. Many FCHVs initially expressed skepticism about the efficacy of pharmacological approaches, reflecting a broader belief that these methods might not be effective or accessible in their context. Increased availability and access to both NRTs and non-NRTs could enhance cessation efforts.

Progress in implementing tobacco cessation strategies, particularly in LMIC has been sluggish [18]. Interventions like even a brief advice from physicians and having national quitlines are shown to be effective in high-income settings [19, 20]. Lack of national quitlines and limited numbers of physicians available in rural areas limits the applicability in low-resource settings like Nepal. Integrating community-based cessation efforts via FCHVs with referrals systems to provide intensive counseling or to use pharmacotherapy can be helpful in LMIC [19, 20].

In Vietnam, at a 6-month follow-up, abstinence rates among smokers were significantly higher when cessation services were provided by both healthcare providers and village health workers compared to delivery solely by healthcare providers (25.7% vs. 10.5%) [21]. Similarly, a study of tobacco cessation program by CHW in Thailand showed the numbers of cigarettes smoked per day reduced by 7.2 with 29% achieving complete cessation at one year [22]. Despite absence of such studies in Nepal, results from other LMICs, together with our findings of FCHV’s potential to deliver tobacco cessation, underscore FCHVs’ potential in tobacco cessation services. If FCHVs can demonstrate effectiveness of tobacco cessation in future clinical trial, they can provide such service in Nepal. Nevertheless, several implementation barriers, including policy level changes are important for FCHVs to successfully deliver and sustain tobacco cessation services.

We believe that our study has implications for research, practice, and policy concerning tobacco cessation interventions in resource-limited settings. Our research adds to the growing evidence supporting the integration of lay health workers like FCHVs to manage chronic diseases. Future research endeavors should continue exploring innovative strategies for maximizing the contribution of FCHVs in tobacco cessation efforts, including developing tailored training programs, appropriate incentives, address specific communities’ needs and challenges.

Our study boasts several strengths. We used a two-phase evaluation following 11 months allowing FCHVs to use the skills they have learned. By sampling FCHVs with diverse backgrounds and integrating both quantitative and qualitative inquiries, we were able to triangulate and enrich our findings. Despite using multiple surveys to track changes, we did not observe major changes. This may be attributed to the FCHVs’ higher baseline knowledge and confidence. However, integrating qualitative data provided a nuanced and in-depth view of complex phenomenon and highlighted areas for improvement, enhancing our understanding of how CHWs can be effectively engaged in tobacco cessation efforts in rural settings.

Limitations of our study include the potential for social desirability bias in self-reported data, and a smaller sample for quantitative evaluations. Although we discussed about motivational interviewing and shared decision making in our trainings, we did not evaluate FCHVs’ competency in these principles in a standardized way. Future work should consider standard evaluation of these principles.

## Conclusion

FCHVs are motivated and have a potential to provide tobacco cessation counseling in rural Nepal. They exhibited confidence and knowledge about tobacco products, its effects on health and society, strategies and challenges for cessation. Future studies should consider addressing identified barriers related to training needs, easy tobacco access, community’s receptivity to cessation counseling and other implementation challenges. Policymakers can consider integration of FCHVs’ tobacco cessation services into the broader healthcare systems and policies. However, before their wider use, we should first evaluate the clinical effectiveness of FCHVs led tobacco cessation intervention.

## Data Availability

The data underlying this study are available upon reasonable request from the corresponding author.

## List of Abbreviations

CHW: Community Health Worker
FCHV: Female Community Health Volunteer
FGD: Focused Group Discussion
LMIC: Low-middle Income Countries
NCD: Non-Communicable Diseases
NRT: Nicotine replacement Therapy

## Declarations

### Ethics approval and consent to participate

The research activities for this study were carried out in accordance with the Declaration of Helsinki. This study is part of an ongoing NCD Nepal Study [9]. We obtained ethical approval from Nepal Health Research Council (Ref: 2483) and a written informed consent from each FCHV. Before obtaining consent, data collectors informed participants that their participation was voluntary, explained the purpose of the study and the interview, and discussed logistics, the use of audio recording, and de-identification procedures.

### Consent for publication

Not Applicable

### Availability of data and materials

The datasets used and/or analysed during the current study are available from the corresponding author on reasonable request.

### Authors’ contributions

Y.P., O.J., B.C., A.S. and S.B. had full access to all the data in the study and take responsibility for the integrity of the data and the accuracy of the data analysis.

*Concept and design:* S.B., S.S, A.T., A.P., P.G., Y.P.

*Acquisition, analysis, or interpretation of data:* All authors

*Drafting of the manuscript:* O.J., B.C., S.B., B.A., Y.P.

*Critical revision of the manuscript for important intellectual content:* All authors

*Obtained funding:* Not applicable

*Supervision:* Y.P., M.B.

## Acknowledgements

We would like to thank Ms. Madhawi Pokharel for proofreading the manuscript.

## Footnotes

Not Applicable

